# Dual-Metric Energy Analysis of Intracranial EEG: Seizure Onset Zones and Energy-Dominant Regions in Drug-Resistant Epilepsy

**DOI:** 10.64898/2026.05.25.26354017

**Authors:** Ahmet Fatih Atik

## Abstract

**Objective:** To determine whether absolute ictal energy on intracranial EEG identifies brain regions whose epileptogenic involvement is attenuated under existing baseline-normalized, dynamic-systems, and event-based frameworks.

**Approach:** Intracranial EEG from 56 patients (five centers; 21 SEEG, 35 ECoG) was analyzed using the Teager-Kaiser Energy Operator computed as z-scored and raw envelopes; energy-dominant network regions (EDNRs) were defined as electrodes whose raw-energy rank exceeded their z-score rank by at least 2 positions. Hilbert decomposition characterized instantaneous amplitude and frequency.

**Main results:** EDNRs were identified in 51 of 56 patients (91%; mean 3.4). Hilbert decomposition revealed elevated baseline amplitude in EDNRs relative to both non-involved regions (*p* < 0.001) and potential seizure onset zones (PSOZs, the top-ranked electrodes under both metrics; *p* = 0.029), with EDNRs participating in seizure-frequency dynamics comparable to PSOZs (mean ictal frequency shift +3.7 versus +4.1 Hz). EDNR detectability correlated directly with electrode count (Spearman *r* = 0.899, *p* < 0.001) without plateau.

**Significance:** Absolute ictal energy identifies an epileptogenic network component with elevated baseline amplitude attenuated under baseline-normalized metrics. The dual-metric framework defines a complementary energy-based axis and establishes the second layer of a two-layer approach with seizure onset and propagation mapping as the first layer. EDNR detectability scales with electrode count, directly relevant to SEEG implantation strategy and to network-level inferences from heterogeneously covered cohorts.

**Highlights:**

- Dual-metric Teager-Kaiser energy operator (TKEO) identifies seizure onset zones and energy-dominant network regions
- Energy-dominant network regions (EDNRs) were identified in 91% of 56 patients across five epilepsy centers
- EDNRs combine highest baseline amplitude with seizure-frequency dynamics; detection scales with electrode count

## 1. Introduction

Approximately one-third of patients with epilepsy have seizures refractory to antiseizure medications, and for these patients, surgical treatment (resection, laser ablation, or neuromodulation) may offer meaningful seizure reduction or freedom (de Tisi et al., 2011; Engel, 1996; Jehi et al., 2015; Kwan and Brodie, 2000). Accurate delineation of the seizure onset zone and the broader epileptogenic network is central to surgical planning (Rosenow and Luders, 2001; Spencer, 2002). Intracranial electroencephalography (iEEG), including stereoelectroencephalography (SEEG) and electrocorticography (ECoG), remains the gold standard for localizing seizure generators prior to intervention, and a substantial quantitative literature has developed over the past two decades to extract interpretable markers of epileptogenicity from these recordings. Yet identifying *which regions of the brain are meaningfully participating in a given patient’s seizures* remains a difficult question in individual cases, and the space of quantitative strategies for interrogating iEEG recordings may not yet be fully mapped.

Three families of quantitative approaches currently dominate intracranial EEG analysis, and each embeds a distinct assumption about how epileptogenic tissue should reveal itself. Baseline-normalized energy measures, exemplified by the Epileptogenicity Index (Bartolomei et al., 2008), express ictal activity as a change from each channel’s own pre-ictal resting state and implicitly assume that epileptogenic regions depart substantially from their own baseline. Dynamic-systems approaches such as neural fragility (Bernabei et al., 2023; Li et al., 2021) characterize seizure generators through the instability they introduce into a linear model of the intracranial network and reference ictal behavior to a resting system configuration. Event-based markers, most prominently high-frequency oscillations (Frauscher et al., 2017; Lisgaras et al., 2026), identify epileptogenic tissue through discrete transient phenomena counted against a background rate. Although conceptually distinct, all three families either normalize ictal activity to a resting reference or distill it into transient events; none directly interrogate the absolute ictal energy that a region sustains throughout a seizure.

This shared design choice may leave a portion of the quantitative picture underexplored. Rosenow and Lüders described the irritative zone as chronically hyperexcitable tissue extending beyond the seizure onset zone (Rosenow and Luders, 2001), and such tissue may plausibly carry elevated baseline activity. A region within the irritative zone that participates strongly in seizures could nonetheless show only a modest *relative* change from its already-elevated resting level, a small frequency shift, and a sparse increment in transient-event rate. Under normalized, stability-based, or event-based metrics, such a region might appear relatively uninvolved, while in absolute terms it could still carry substantial ictal energy throughout the seizure. Whether regions with this profile exist in practice, how prevalent they are, and what their biophysical signature looks like has received limited direct attention.

The present study explores this gap and outlines a two-layer approach to characterizing each patient’s epileptogenic network from SEEG data. The first layer comprises seizure onset zone detection and, when diffusion imaging is available, deterministic tractography-based mapping of the propagation network. The second layer focuses on energy-dominant network regions (EDNRs), regions whose absolute ictal energy is high but whose normalized energy is low, and examines their relationship to the initial network to inform surgical decision-making. A simple operational test is proposed in which each electrode’s rank under a z-scored (normalized) TKEO metric is compared against its rank under a raw (absolute) TKEO metric; EDNRs are defined as regions whose raw-energy rank exceeds their z-score rank. The Teager-Kaiser Energy Operator is used as a minimally-assumption-laden instantaneous energy measure (Kaiser, 1990; Mukhopadhyay and Ray, 1998) rather than as a central methodological contribution. Because the publicly available multicenter datasets used here do not include diffusion tensor imaging, the tractography component of Layer 1 cannot be demonstrated in this cohort. The present study therefore focuses on establishing Layer 2: on a multicenter cohort of 56 patients from two publicly available iEEG datasets, three questions are examined: (i) whether regions with this discordant energy profile can be identified and how prevalent they appear; (ii) what their biophysical signature looks like in terms of instantaneous amplitude and frequency; and (iii) how their detectability varies with electrode sampling density.

## Method

### Study Population

Intracranial EEG data were analyzed from 56 patients with drug-resistant epilepsy across two publicly available multicenter datasets: (1) OpenNeuro ds004100 (Hospital of the University of Pennsylvania [HUP]; n = 26) (Bernabei et al., 2023), and (2) OpenNeuro ds003029 (Johns Hopkins Hospital [JHH], n = 4; National Institutes of Health [NIH], n = 14; University of Michigan Flint [UMF], n = 5; University of Maryland Medical Center [UMMC], n = 7) (Li et al., 2021). Six additional HUP recordings accessed through the iEEG.org portal lacked entries in the ds004100 demographic table and were excluded from all analyses because intracranial monitoring modality (depth versus subdural) could not be confirmed for those patients. Implant modality was resolved from the ds004100_meta participants table for HUP patients and from the BIDS acquisition tag for ds003029 patients, yielding 21 SEEG and 35 ECoG implantations across the 56-patient analysis cohort. Among the 56 patients, the mean age was 36.0 years (range 13-59; n = 52 with age recorded) and 52% were female (27/52 with sex recorded). All patients underwent intracranial monitoring as part of presurgical evaluation, followed by surgical resection or laser ablation. Single-timepoint surgical outcome annotations were available for all 56 patients in the source dataset metadata; these were used only for descriptive analyses as detailed below. Because all data were publicly available and de-identified, institutional review board approval was not required.

### Signal Preprocessing

Raw iEEG signals were converted to a bipolar montage by computing the difference between adjacent contacts on each electrode to reduce common-mode noise and improve spatial specificity. The bipolar signals were bandpass filtered (10-100 Hz) using a fourth-order zero-phase Butterworth filter to isolate the frequency band most relevant to seizure activity while rejecting low-frequency drift and high-frequency noise.

### TKEO Computation and Dual-Metric Quantification

The Teager-Kaiser Energy Operator (Kaiser, 1990; Mukhopadhyay and Ray, 1998) was applied to each bipolar channel. For a discrete signal x[n], the TKEO is defined as:

Psi[x(n)] = x(n)^2 - x(n-1) * x(n+1)

For narrowband signals the TKEO output can take negative values; the absolute value was therefore taken at each sample to yield a non-negative instantaneous energy estimate. The resulting signal was then smoothed using a 20-millisecond moving average window to produce a TKEO envelope for each channel.

**Metric 1: Z-Scored TKEO (Normalized Energy).** A pre-ictal baseline consisting of the first 30 seconds of each recording was used to compute the mean and standard deviation of the TKEO envelope for each channel. For every patient in this cohort, annotated seizure onset occurred at or after 30 seconds into the recording, such that the baseline window preceded ictal activity. Following the baseline normalization approach used for TKEO-based muscle-onset detection (Solnik et al., 2010), the z-scored TKEO was computed as:

z(t) = (TKEO_envelope(t) - mu_baseline) / sigma_baseline

**Metric 2: Raw TKEO (Absolute Energy).** The raw TKEO envelope values were retained in their original units (microvolts squared), capturing the absolute energy burden regardless of baseline levels.

### Onset Detection

Seizure onset was detected per channel from the z-scored TKEO. For each channel, the algorithm identified the first time point at which the z-scored TKEO exceeded the threshold and remained above it for a minimum sustained duration of 100 milliseconds. A per-recording threshold rule was applied to accommodate between-recording amplitude-scale heterogeneity arising from hardware and unit differences across centers: z = 200 when the peak z-score exceeded 500, and z = 50 otherwise.

### EDNR Identification

For each electrode (corresponding to an anatomical brain region), peak z-scored TKEO and peak raw TKEO values were computed. All electrodes were ranked independently by each metric (rank 1 = highest). An electrode was classified as an energy-dominant network region (EDNR) when its z-score rank minus its raw-energy rank was 2 or greater, indicating that the electrode appeared at least two positions more prominent under the raw (absolute) metric than under the normalized metric. To assess robustness of this operational definition, sensitivity analyses were performed in which the rank-discordance threshold was varied between 1 and 3 positions. To assess whether findings were driven by the inclusion of one monitoring modality, the principal analyses (prevalence, electrode-coverage scaling, and descriptive clinical associations) were repeated separately in the SEEG (n = 21) and ECoG (n = 35) subcohorts.

### Amplitude-Frequency Decomposition

To investigate the biophysical signature of EDNRs, the bandpass-filtered bipolar signal was decomposed into instantaneous amplitude and instantaneous frequency components via the Hilbert transform. For each bipolar channel, the analytic signal was computed to yield the instantaneous amplitude envelope and instantaneous frequency time series. The following electrode-level metrics were then computed: (1) the amplitude ratio, defined as peak ictal amplitude divided by mean pre-ictal amplitude, yielding a dimensionless amplification factor; (2) the frequency shift, defined as mean ictal frequency minus mean pre-ictal frequency, in Hz; and (3) the baseline amplitude, defined as the mean pre-ictal amplitude envelope in microvolts. Each electrode was assigned to one of three classes derived from the dual-metric analysis: EDNR (as defined above); potential seizure onset zone (PSOZ), defined operationally as electrodes ranked at the top under both the normalized and raw metrics and used here as the algorithmic SOZ candidates output by the dual-metric pipeline; or non-involved (electrodes ranked low under both metrics). The PSOZ designation is intentionally labeled “potential” because clinical SOZ annotations are not uniformly available across the public datasets used here, and the dual-metric pipeline identifies these regions algorithmically rather than confirming them against gold-standard clinical annotation. Group comparisons were performed using the two-tailed Mann-Whitney U test. Four patients (umf002, umf003, umf004, umf005) were excluded from this analysis because their publicly available recording clips were 0.7-2.0 seconds in duration, insufficient to separate pre-ictal and ictal windows; Hilbert decomposition was therefore performed on the remaining 52 patients of the analysis cohort.

### Electrode Coverage Analysis

The relationship between electrode sampling density and EDNR detectability was assessed with Spearman rank correlation between the number of electrodes per patient and the number of EDNRs identified. Patients were stratified into low (3-10 electrodes), medium (11-14 electrodes), and high (15 or more electrodes) coverage tiers. Cut-points were chosen to align with clinically meaningful SEEG coverage levels observed across the cohort. The proportion of sampled regions classified as EDNRs was computed for each tier.

### Anatomical Classification

For all 26 HUP patients in the analysis cohort (for whom MNI-space electrode coordinates were publicly available), each SEEG or ECoG contact was assigned to an anatomical parcel using the Harvard-Oxford cortical and subcortical maximum-probability atlas at 1 mm resolution (no probability threshold). Within each electrode, the majority parcel across contacts was taken as the electrode-level anatomical assignment. Parcels were grouped into major lobe categories (frontal, temporal, mesial temporal, parietal, occipital, insular, cingulate). Electrode coordinates were not included in the ds003029 public release, and anatomical classification could not be performed for those patients.

### Descriptive Clinical Annotations

For all 56 patients in the analysis cohort, the source datasets included single-timepoint outcome annotations using the Engel classification (Engel et al., 1993). Outcome harmonization across the two source datasets was possible only at the binary level: ds004100 reported sub-classed Engel scores (1A through 4A), therapy type, surgical target, and lesion status but not follow-up duration; ds003029 reported integer-level Engel scores, the ILAE outcome scale, and post-operative follow-up duration but not therapy type or lesion status. EDNR counts and the presence of bilateral EDNRs were therefore compared at the binary level between patients labeled seizure-free (Engel I) and not seizure-free (Engel II-IV) using two-tailed Mann-Whitney U and Fisher exact tests, respectively. These comparisons are reported as descriptive observations only (see Limitations).

### Statistical Analysis

All statistical analyses were performed in Python 3.13 using SciPy 1.14 and statsmodels 0.14. Two-tailed Mann-Whitney U tests were used for all between-group comparisons of continuous variables because the data were not normally distributed. Fisher’s exact test was used for comparison of categorical variables. Spearman rank correlation was used to assess the monotonic relationship between electrode coverage and EDNR count. *P* values are reported to two decimal places when greater than 0.01 and to three decimal places when between 0.001 and 0.01; values less than 0.001 are reported as *p* < 0.001. No correction for multiple comparisons was applied given the exploratory nature of these analyses. This study conforms to the STROBE guidelines for reporting observational studies.

## 2. Results

### 2.1 Patient Characteristics

The analysis cohort comprised 56 patients from five epilepsy centers (Table 1). Intracranial monitoring was performed with SEEG in 21 patients and ECoG in 35. Electrode coverage ranged from 3 to 26 electrodes per patient (mean 11.8, median 12), yielding a mean of 98 bipolar channels per patient (range 32-204).

**Table 1.**
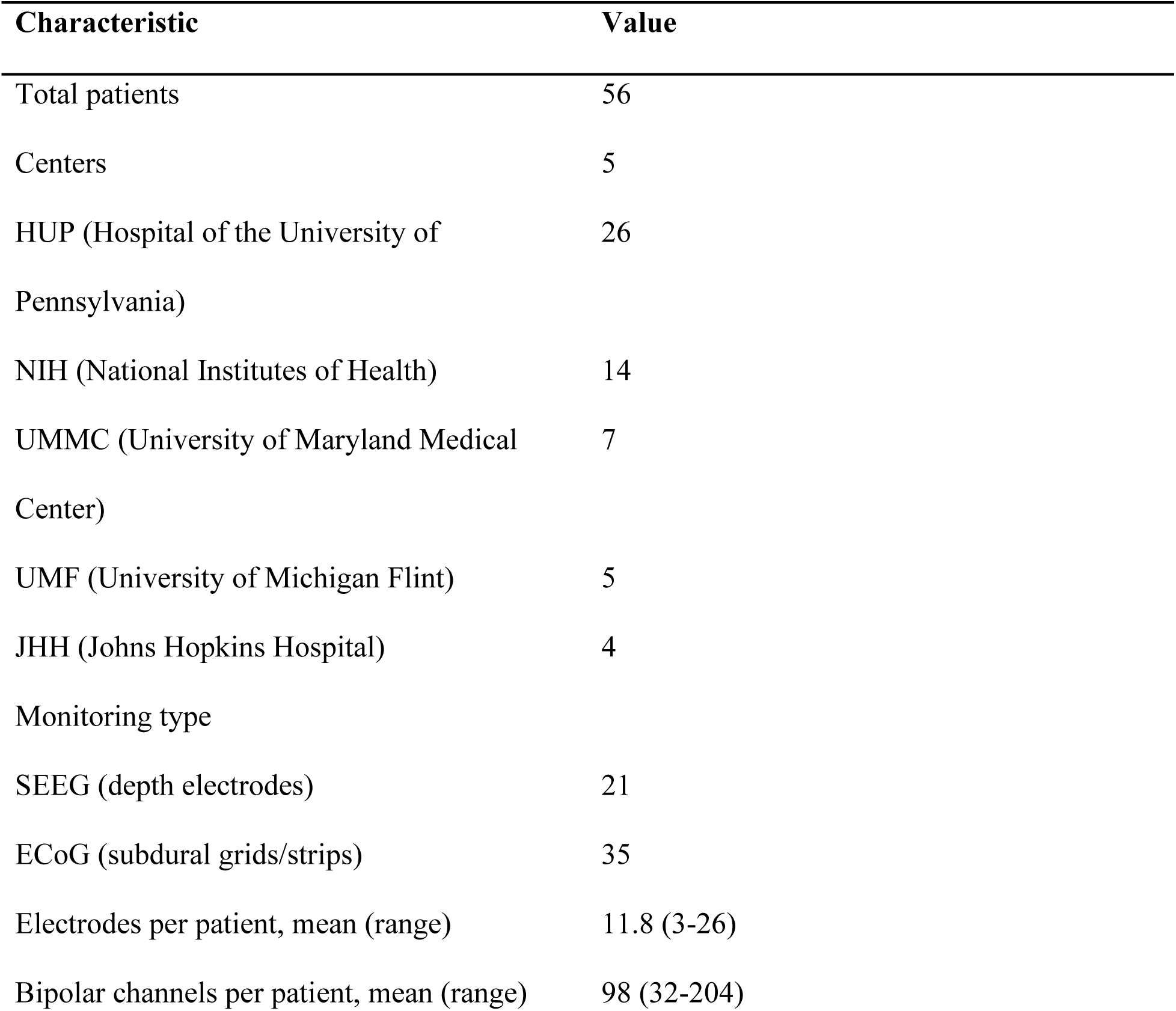
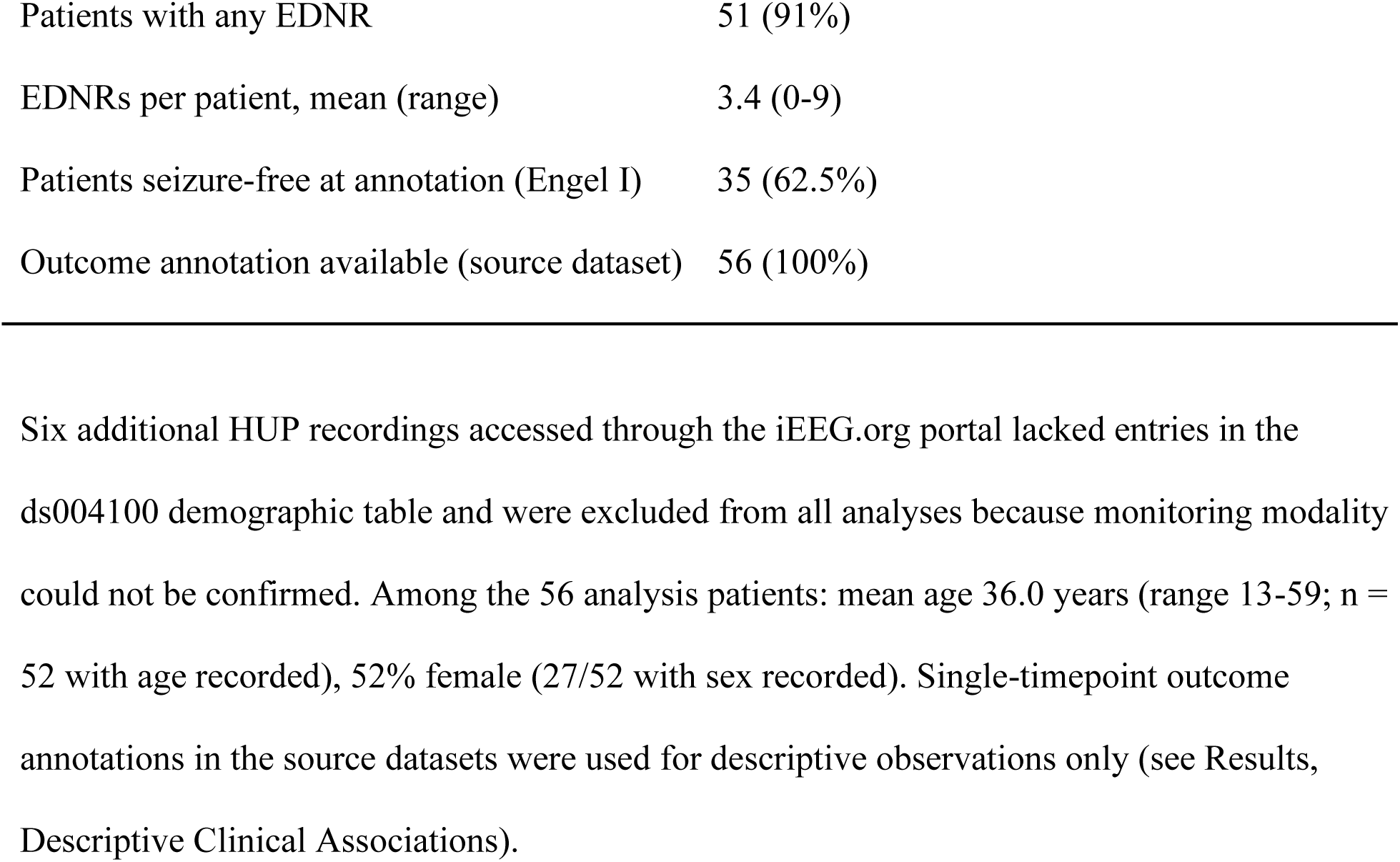
Cohort characteristics. Six additional HUP recordings accessed through the iEEG.org portal lacked entries in the ds004100 demographic table and were excluded from all analyses because monitoring modality could not be confirmed. Among the 56 analysis patients: mean age 36.0 years (range 13-59; n = 52 with age recorded), 52% female (27/52 with sex recorded). Single-timepoint outcome annotations in the source datasets were used for descriptive observations only (see Results, Descriptive Clinical Associations).

### 2.2 Outcome Reporting Across Source Datasets

The two source datasets reported surgical outcome with markedly different granularity (Table 2). ds004100 provided Engel sub-classes (eight categories: 1A, 1B, 1C, 1D, 2A, 2C, 3A, 4A), therapy type (ablation versus resection), surgical target, and lesion status, but did not record post-operative follow-up duration. ds003029 provided integer-level Engel score (no sub-classification), the ILAE outcome scale, and follow-up duration, but did not record therapy type, surgical target, or lesion status. The only outcome variable that could be harmonized across both datasets was a binary classification of seizure-free (Engel I) versus not seizure-free (Engel II-IV). In the 56-patient analysis cohort, 35 patients (62.5%) were classified as seizure-free at the time of dataset annotation. Median post-operative follow-up duration in ds003029 was 1 year (range 1-7 years); follow-up timing was not recorded in ds004100. Because longer post-operative surveillance generally reveals late recurrences and reduces apparent seizure-freedom rates relative to short-term assessment (de Tisi et al., 2011), the 62.5% seizure-free rate observed in this cohort represents an upper bound on the durable seizure-freedom rate that would be observed in a longer-follow-up sample of the same population.

**Table 2.**
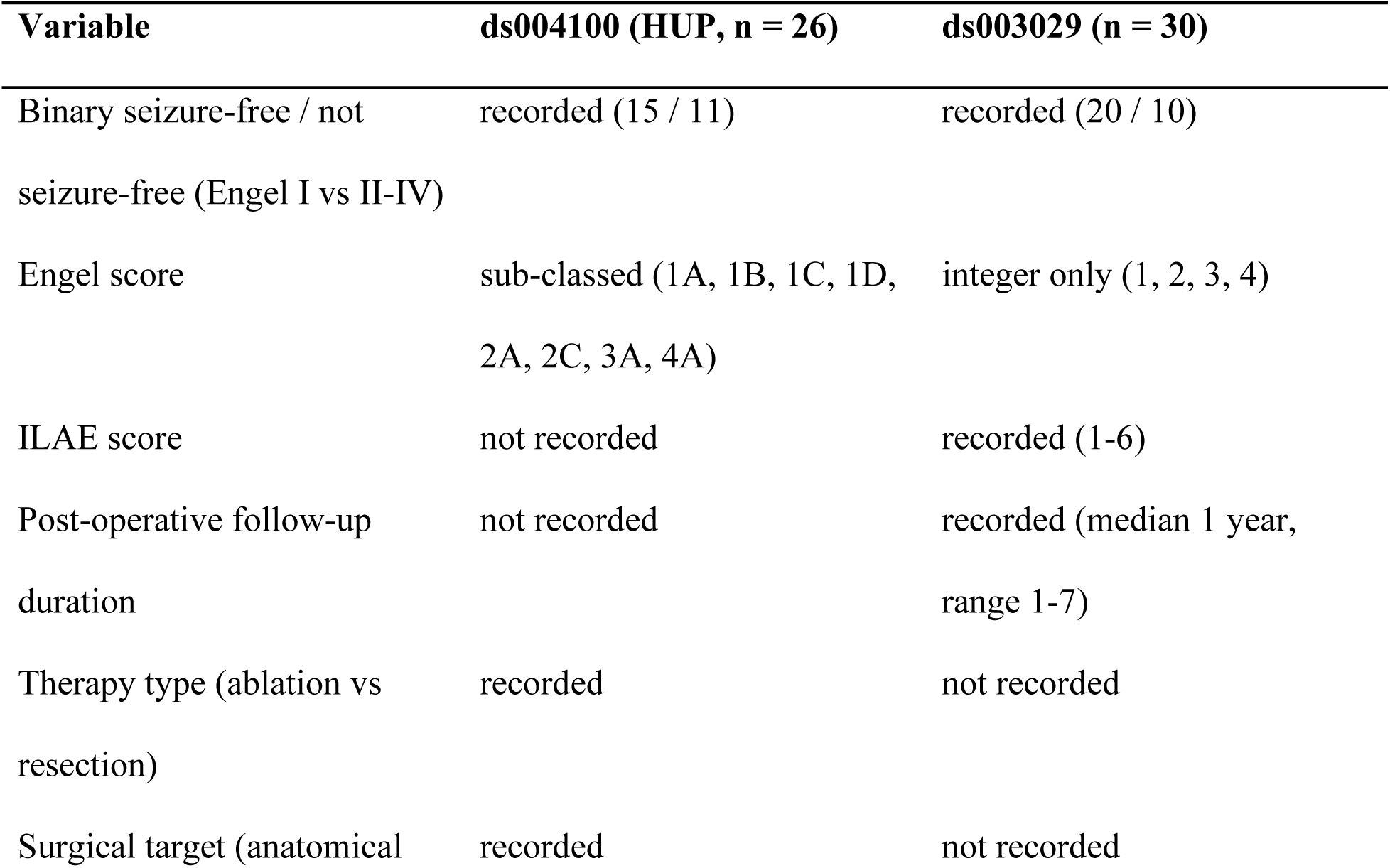
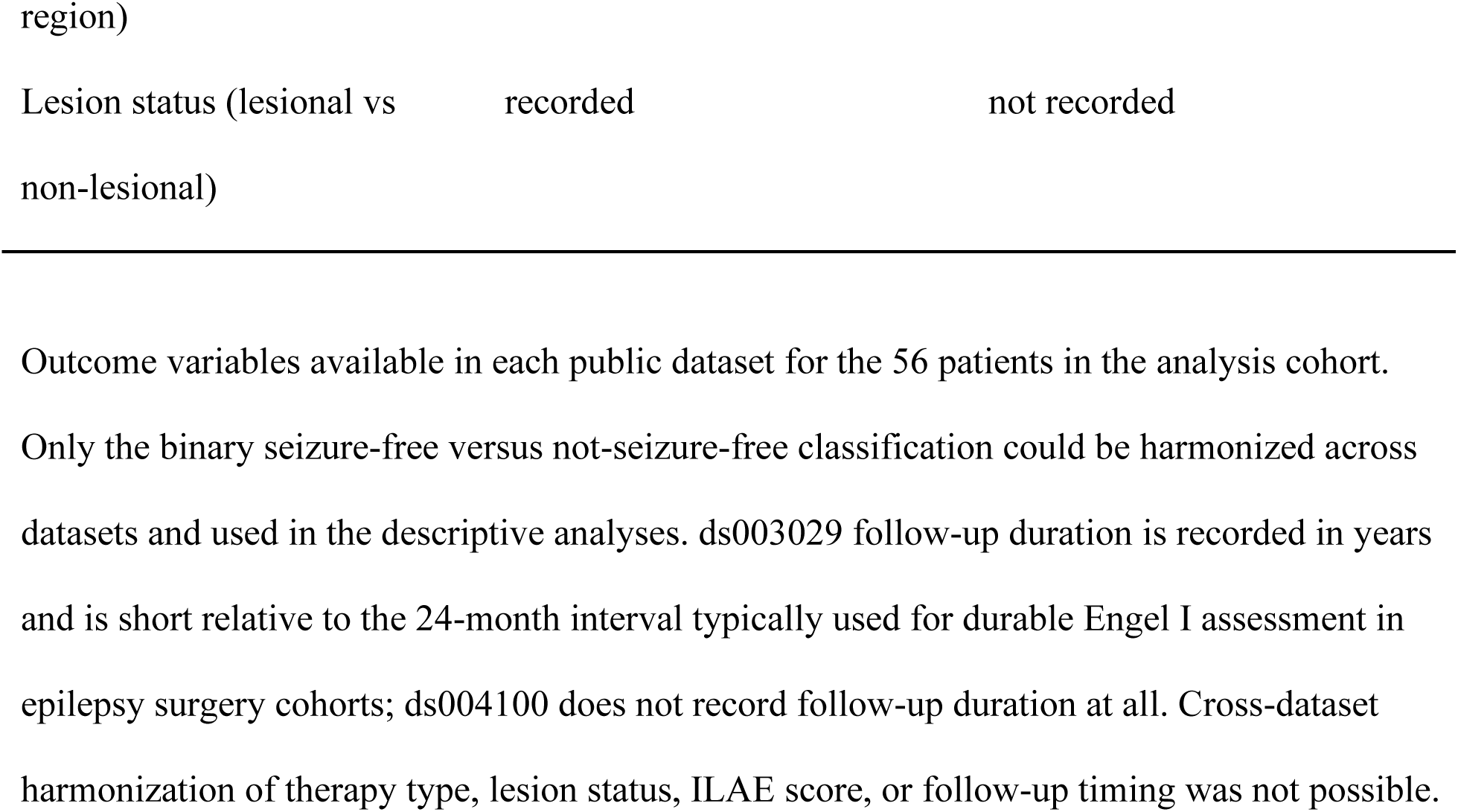
Outcome reporting variables across source datasets. Outcome variables available in each public dataset for the 56 patients in the analysis cohort. Only the binary seizure-free versus not-seizure-free classification could be harmonized across datasets and used in the descriptive analyses. ds003029 follow-up duration is recorded in years and is short relative to the 24-month interval typically used for durable Engel I assessment in epilepsy surgery cohorts; ds004100 does not record follow-up duration at all. Cross-dataset harmonization of therapy type, lesion status, ILAE score, or follow-up timing was not possible.

### 2.3 Identification and Prevalence of Energy-Dominant Network Regions

Dual-metric analysis revealed a consistent pattern across the cohort. In 51 of 56 patients, a subset of brain regions ranked highly by raw TKEO energy but poorly by z-scored TKEO energy, a pattern compatible with chronically elevated baseline activity whose seizure involvement may be attenuated under normalization. These discordant regions (energy-dominant network regions; EDNRs) were identified in 51 of 56 patients (91%; Table 1), with a mean of 3.4 per patient (range 0-9). Only 5 of 56 patients had no EDNRs. The distribution of rank discordance scores across all identified EDNRs and the per-patient EDNR count distribution are shown in Figure 1A and Figure 1B, respectively. Figure 2 illustrates the dual-metric comparison in an exemplar patient (HUP158), in which regions that rank low by z-score rank high by raw energy; this pattern is not apparent under any single-metric analysis.

**Figure 1.**
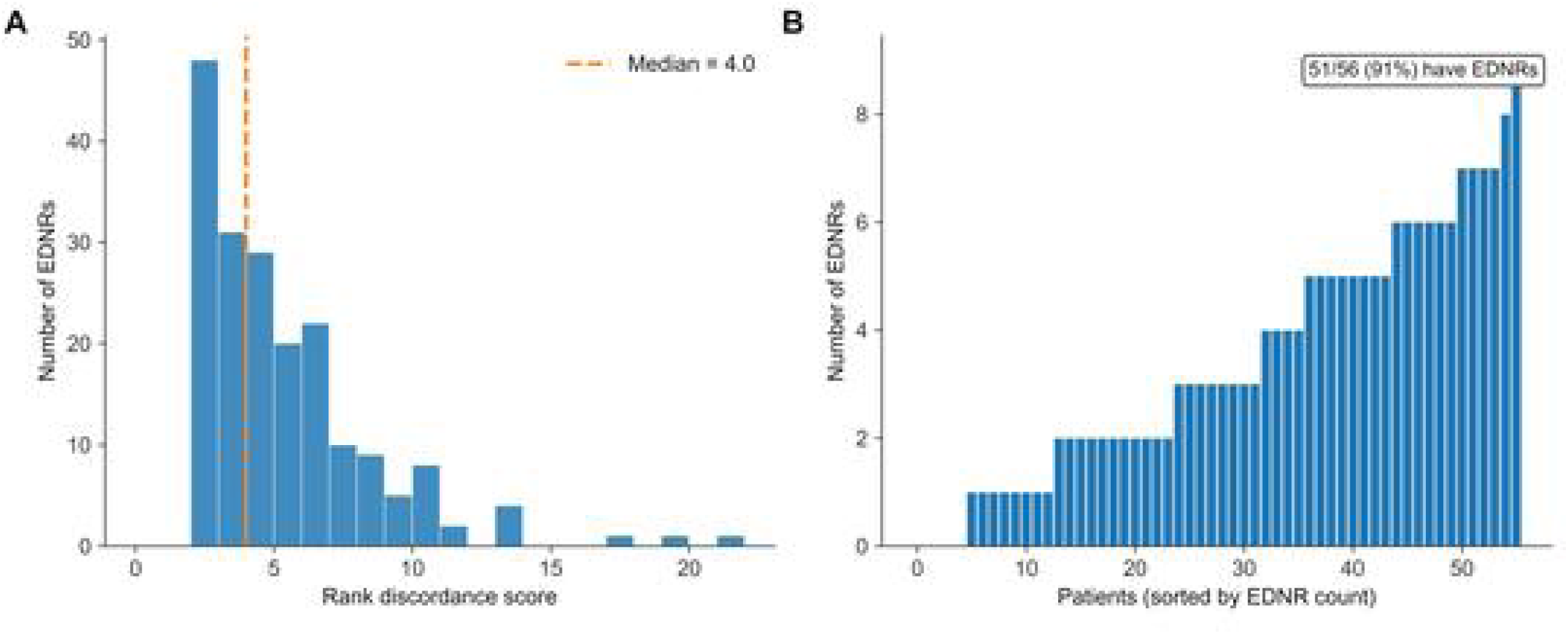
Distribution of rank discordance and EDNR prevalence across the cohort. (**A**) Histogram of rank discordance scores for all EDNRs identified across the 56 patients in the analysis cohort. Rank discordance is defined as the absolute difference between a region’s z-scored energy rank and its raw energy rank; higher values indicate greater disagreement between normalized and absolute metrics. Dashed line indicates the median discordance score. (**B**) Number of EDNRs per patient, sorted by count. Each horizontal bar represents one patient. EDNRs were identified in 51 of 56 patients (91%) with a mean of 3.4 per patient (range 0-9).

**Figure 2.**
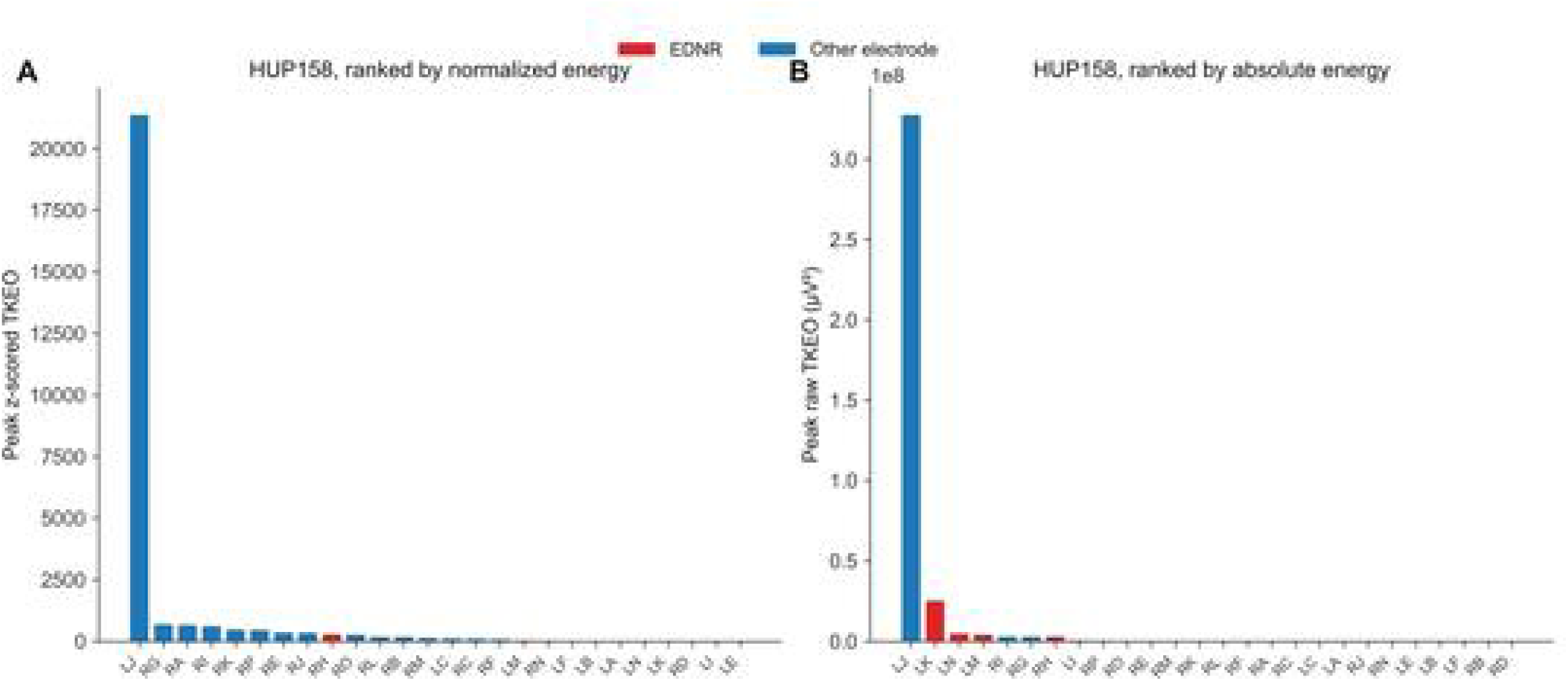
Dual-metric energy comparison in an exemplar patient (HUP158). (**A**) All electrodes ranked by peak z-scored TKEO energy. Blue bars indicate potential seizure onset zones (PSOZs), the regions ranked at the top under the normalized metric and corresponding to the algorithmic SOZ candidates output by the dual-metric pipeline. Under this normalized metric, several regions appear minimally involved. (**B**) The same electrodes ranked by peak raw (absolute) TKEO energy in microvolts squared. Red bars indicate EDNRs, regions that rank low by z-score (panel A) but high by raw energy (panel B), revealing substantial absolute ictal energy burden that is not apparent under normalized detection. The discordance between the two rankings illustrates how baseline normalization structurally attenuates the contribution of regions with chronically elevated baseline activity.

Anatomical distribution of EDNRs was characterized in the subset of 26 HUP patients for whom publicly available electrode coordinates permitted atlas-based classification (Figure 4). EDNRs were identified across every sampled lobe, with frontal (n = 31), temporal (n = 20), and mesial temporal (n = 16) regions contributing the largest absolute counts. When adjusted for the number of electrodes sampled in each lobe, per-electrode EDNR rates were broadly comparable across frontal (31%), mesial temporal (31%), parietal (27%), and temporal (22%) regions, suggesting that EDNRs are not restricted to a specific anatomical territory. Electrode coordinates were not included in the ds003029 public release, precluding anatomical classification for those patients. Sensitivity analyses across rank-discordance thresholds of 1, 2, and 3 positions yielded qualitatively consistent findings. Cohort prevalence was 98%, 91%, and 82% and mean EDNRs per patient were 4.5, 3.4, and 2.6 at thresholds of ≥ 1, ≥ 2, and ≥ 3, respectively. The directional outcome and bilateral-involvement patterns reported below were preserved at every threshold.

### 2.4 Biophysical Signature of EDNRs

To characterize the biophysical profile of EDNRs, the bandpass-filtered bipolar signal was decomposed into instantaneous amplitude and frequency components via the Hilbert transform in 52 patients with sufficient data quality (Table 3). The central observation is that EDNRs carry the highest baseline amplitude of any region class in the cohort. EDNR baseline amplitude was significantly elevated relative to both non-involved regions (two-tailed Mann-Whitney U, *p* < 0.001) and PSOZs (*p* = 0.029), with PSOZs themselves showing baseline amplitudes elevated above non-involved regions. This elevated baseline is the mechanism proposed in the Introduction made concrete: a large z-score denominator scales with chronic baseline activity, so the same absolute ictal energy that would produce a high z-score in a quiet region produces only a modest z-score in a region with chronically elevated baseline.

**Table 3.**
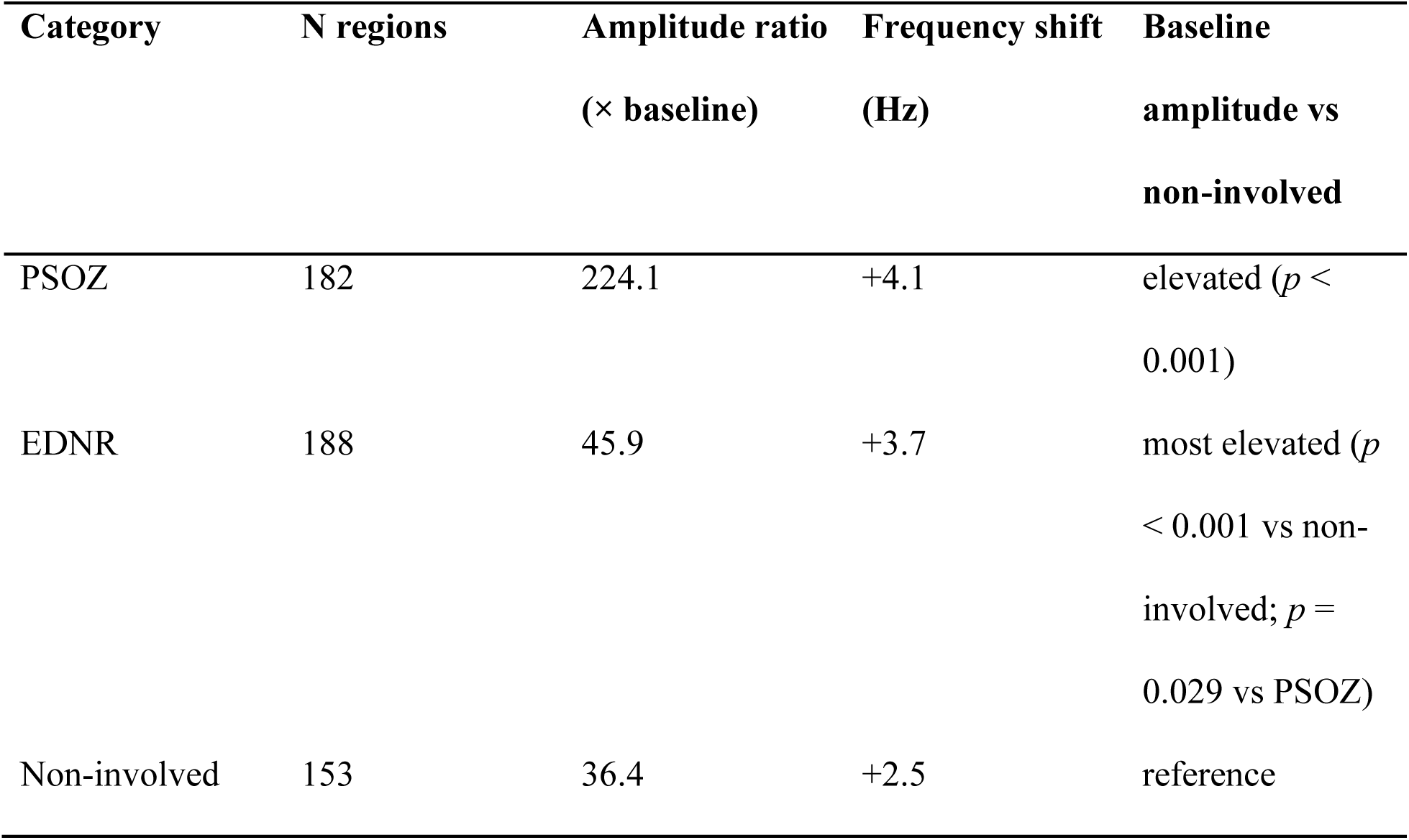
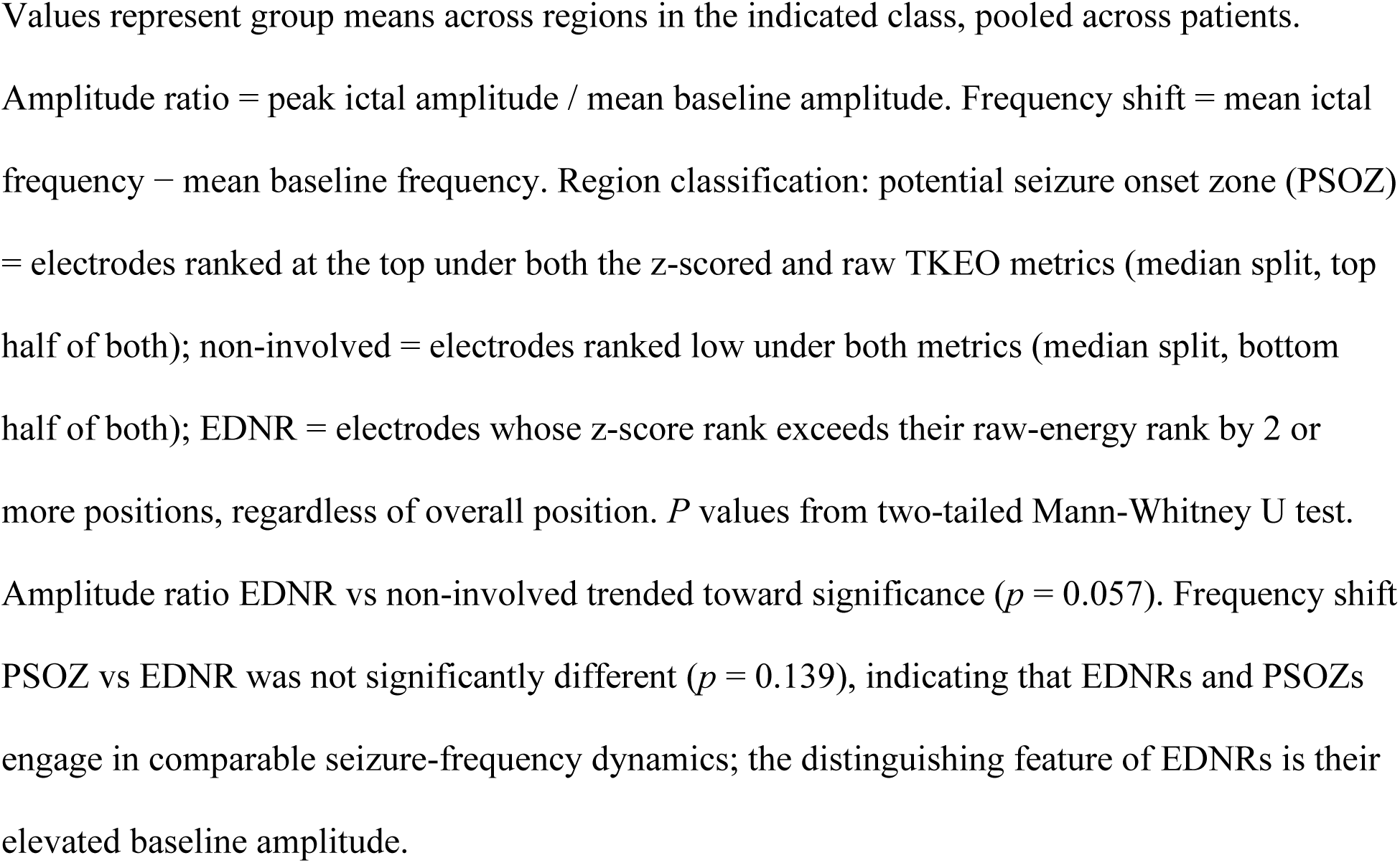
Amplitude-frequency decomposition by region classification (n = 52). Values represent group means across regions in the indicated class, pooled across patients. Amplitude ratio = peak ictal amplitude / mean baseline amplitude. Frequency shift = mean ictal frequency − mean baseline frequency. Region classification: potential seizure onset zone (PSOZ) = electrodes ranked at the top under both the z-scored and raw TKEO metrics (median split, top half of both); non-involved = electrodes ranked low under both metrics (median split, bottom half of both); EDNR = electrodes whose z-score rank exceeds their raw-energy rank by 2 or more positions, regardless of overall position. *P* values from two-tailed Mann-Whitney U test. Amplitude ratio EDNR vs non-involved trended toward significance (*p* = 0.057). Frequency shift PSOZ vs EDNR was not significantly different (*p* = 0.139), indicating that EDNRs and PSOZs engage in comparable seizure-frequency dynamics; the distinguishing feature of EDNRs is their elevated baseline amplitude.

Two further observations support the interpretation that EDNRs are genuine seizure participants rather than passive bystanders. First, EDNRs showed substantial ictal amplitude amplification (mean 45.9-fold over baseline), trending above the 36.4-fold amplification observed in non-involved regions (two-tailed *p* = 0.057), and far below the 224-fold amplification observed in PSOZs. Second, EDNRs showed a mean ictal frequency shift of +3.7 Hz, comparable to the +4.1 Hz shift in PSOZs and well above the +2.5 Hz shift in non-involved regions. EDNRs therefore engage in seizure-frequency dynamics in a manner similar to PSOZs, indicating active ictal participation rather than passive volume conduction. The distinguishing feature of EDNRs relative to PSOZs is their elevated baseline, not a different ictal profile.

Taken together, these observations describe a physiological profile in which EDNRs participate in seizure activity comparably to PSOZs but enter that activity from a substantially higher baseline. The elevated baseline alone is sufficient to explain why EDNRs may produce attenuated signals under metrics that reference ictal activity to a resting baseline (z-score normalization, Epileptogenicity Index), to a resting network configuration (neural fragility), or to a discrete background rate (high-frequency oscillations).

### 2.5 Scaling of EDNR Detection with Electrode Coverage

The number of EDNRs identified per patient increased strongly with electrode sampling density. The Spearman correlation between electrode count and EDNR count was *r* = 0.899 (*p* < 0.001; Figure 3A). Stratifying patients by coverage tier showed that the proportion of sampled regions classified as EDNRs rose from 20.2% in patients with low coverage (3-10 electrodes) to 27.4% at medium coverage (11-14 electrodes) and 35.7% in patients with high coverage (15 or more electrodes); the difference across tiers was significant on chi-square testing (*p* = 0.001; Figure 3B). No evidence of saturation was observed within the coverage range present in these datasets, though the maximum of 26 electrodes per patient may be insufficient to detect a plateau if one exists at higher sampling densities.

**Figure 3.**
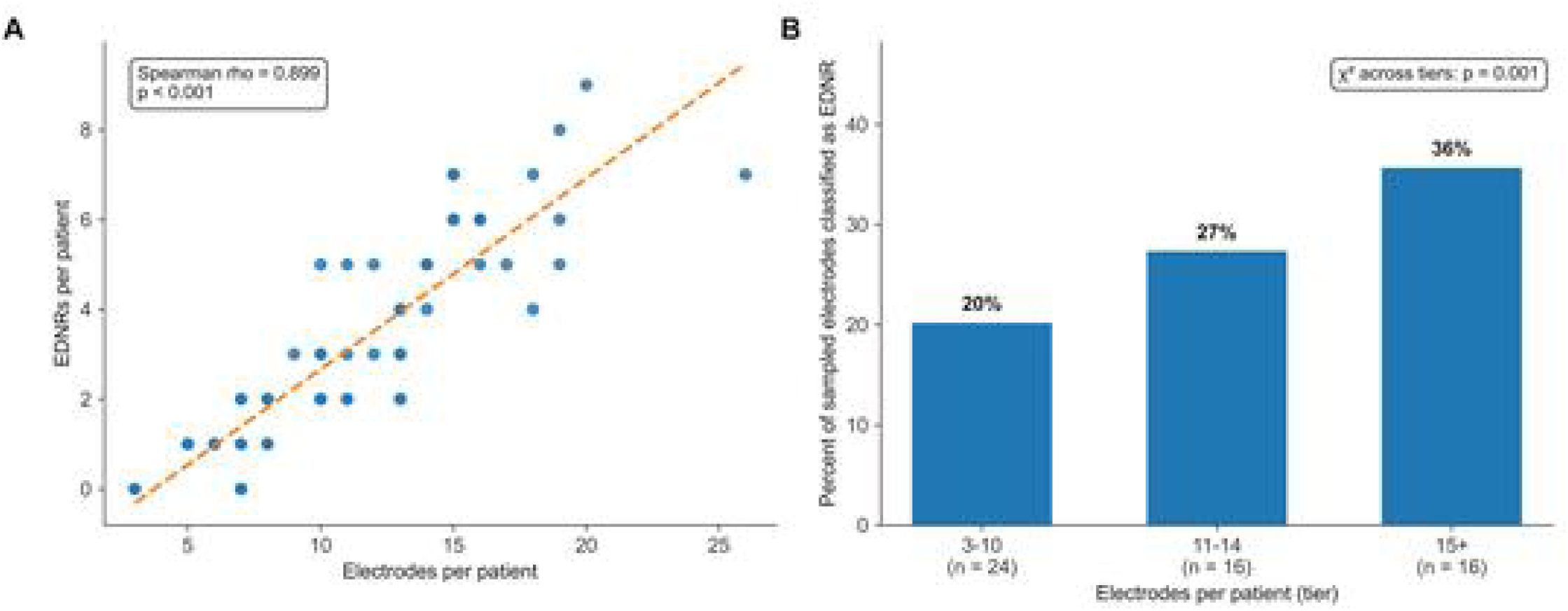
Scaling of EDNR detection with electrode sampling density. (**A**) Scatter plot of number of electrodes per patient versus number of EDNRs detected (Spearman *r* = 0.899, *p* < 0.001). Each dot represents one patient. The strong positive correlation without any apparent plateau indicates that EDNR detection continues to increase with sampling density across the full coverage range present in these datasets. (**B**) Proportion of sampled regions classified as EDNRs by electrode coverage tier: 20.2% in patients with low coverage (3-10 electrodes), 27.4% at medium coverage (11-14 electrodes), and 35.7% at high coverage (15 or more electrodes); χ² test across tiers *p* = 0.001.

This scaling has two interpretations, and both carry implications for SEEG practice. The first is that EDNRs are a distributed physiological population: their true extent exceeds what any single electrode array fully samples, and denser implantations reveal progressively more of the network. The second is that any comparison of EDNR counts across patients with heterogeneous coverage is inherently conditioned on sampling density; cross-patient comparisons on mixed-coverage cohorts cannot be interpreted as unbiased estimates of underlying network burden. The first interpretation is a finding about the epileptogenic network; the second is a methodological constraint on any study (including this one) that analyzes public multicenter iEEG data with heterogeneous implantation strategies.

### 2.6 Descriptive Clinical Associations

In the 56-patient analysis cohort, the number of EDNRs per patient appeared directionally higher in patients labeled not seizure-free than in patients labeled seizure-free (mean 4.14 vs. 2.97; median 3 vs. 2; two-tailed Mann-Whitney *p* = 0.083). A monotonic gradient across annotated Engel classes was also observed (median EDNRs: I = 2, II = 3, III = 5, IV = 3; with the small Engel IV subgroup [n = 5] limiting interpretation of the highest class). Bilateral EDNRs appeared more frequent in the not-seizure-free group than in the seizure-free group (48% vs. 20%; odds ratio 3.64; two-tailed Fisher exact *p* = 0.039).

These observations are offered here as descriptive only. The outcome fields in the public datasets are single-timepoint annotations without longitudinal follow-up, chart review, harmonized scoring intervals, or knowledge of resection volumes, and cannot support any claim of predictive validity. They are noted because their direction may be consistent with the possibility that EDNRs carry some clinical relevance, a possibility that would need to be tested in prospective cohorts with standardized coverage and longitudinal surveillance (see Discussion).

### 2.7 Sensitivity to Monitoring Modality

To address whether the principal findings reflected the inclusion of subdural-grid (ECoG) monitoring rather than depth-electrode (SEEG) recordings, the analyses were repeated separately in the SEEG (n = 21) and ECoG (n = 35) subcohorts. EDNRs were identified in 21 of 21 SEEG patients (100%; mean 4.5 per patient) and in 30 of 35 ECoG patients (86%; mean 2.7 per patient), with electrode-coverage scaling preserved in both subgroups (Spearman *r* = 0.815 for SEEG, *r* = 0.877 for ECoG; both *p* < 0.001). Bilateral EDNRs were directionally more frequent in not-seizure-free than in seizure-free patients within both subcohorts (SEEG: 70% vs. 45%, Fisher OR = 2.80, *p* = 0.387; ECoG: 27% vs. 8%, Fisher OR = 4.12, *p* = 0.297). The reduction in statistical significance relative to the full-cohort association (OR = 3.64, *p* = 0.039) is consistent with the expected loss of power when the cohort is divided rather than absence of effect, and the subgroup odds ratios bracket the full-cohort estimate.

Two observations follow. First, the higher per-patient EDNR yield in SEEG (4.5 versus 2.7) and the higher prevalence (100% versus 86%) are consistent with depth electrodes’ anatomical reach into brain regions that subdural grids cannot sample, including mesial temporal structures, the insula, and the depths of sulci, where many EDNRs in this cohort were identified (Figure 4). Second, the comparable Fisher odds ratios in both subgroups indicate that the EDNRs that ECoG does identify carry the same association with outcome as those that SEEG identifies. ECoG appears to under-sample EDNRs through its inherent geometric limitation rather than to identify a structurally different signal, suggesting that the modalities differ in the volume of brain accessible to monitoring rather than in what they detect when sampling is possible.

**Figure 4.**
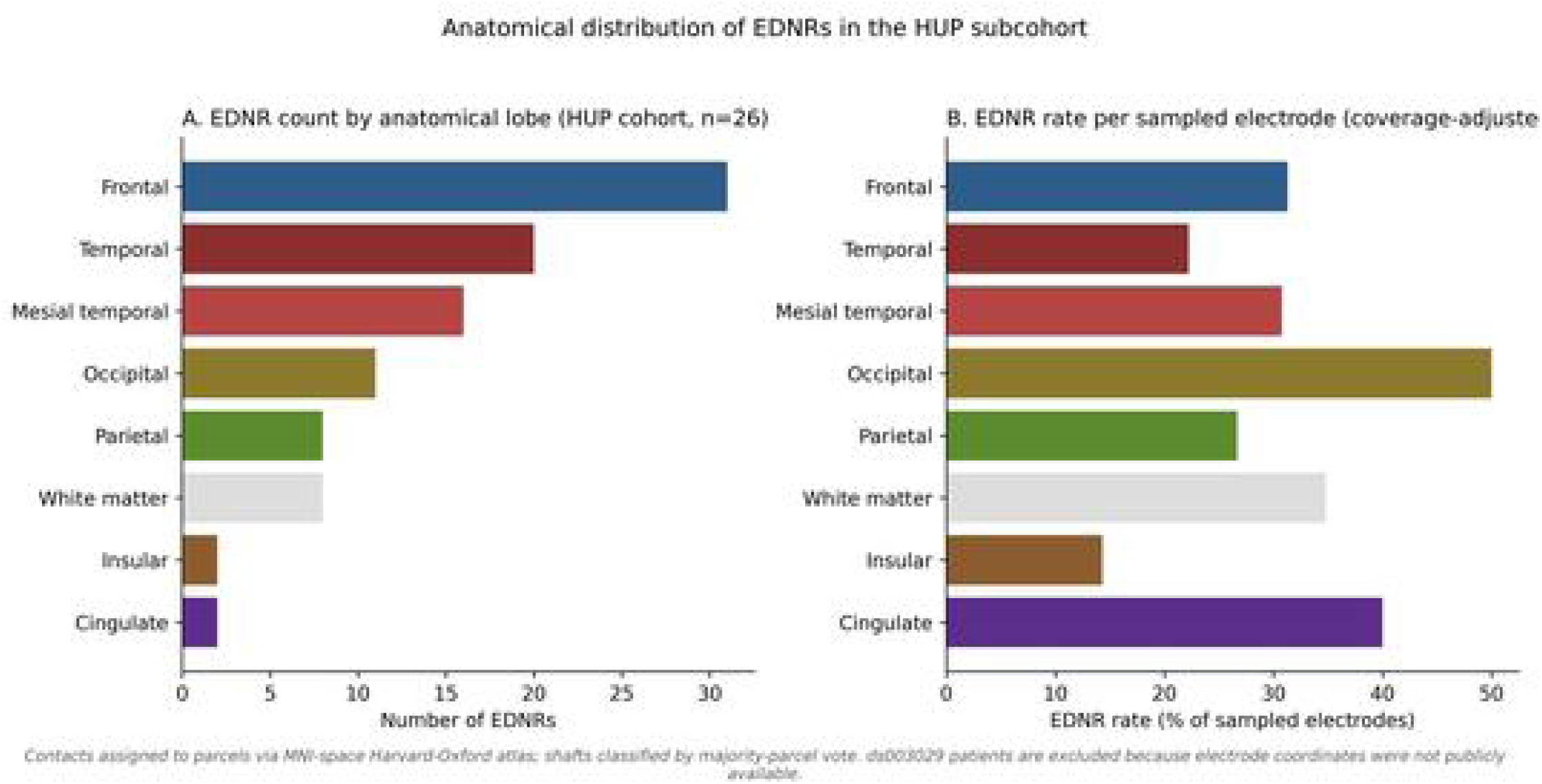
Anatomical distribution of EDNRs in the HUP subcohort (n = 26 patients with publicly available electrode coordinates). Each electrode contact was assigned to a parcel using the MNI-space Harvard-Oxford cortical and subcortical atlas; electrode-level anatomical assignment was determined by majority-parcel vote across contacts. (**A**) Absolute EDNR count by anatomical lobe. Frontal (n = 31), temporal (n = 20), and mesial temporal (n = 16) regions contributed the largest absolute counts, reflecting both EDNR prevalence and the distribution of electrode sampling across lobes. (**B**) Per-electrode EDNR rate (percentage of sampled electrodes classified as EDNRs) by anatomical lobe. Rates were broadly comparable across lobes, suggesting that EDNR occurrence is not restricted to a specific anatomical territory. ds003029 patients were excluded because electrode coordinates were not included in the public release.

## 3. Discussion

### 3.1 A Physiologically Distinct Component of the Epileptogenic Network

The present analysis describes a physiologically distinct component of the epileptogenic network that may not be fully captured by several of the quantitative frameworks currently used in intracranial EEG analysis. The findings define a previously uncharacterized dimension of epileptogenic network organization and establish the second layer of a two-layer clinical framework. Rather than proposing a new biomarker in isolation, they draw attention to a structural assumption shared by existing methods and supply a complementary energy-based axis that, when paired with seizure onset detection and tractography-based propagation mapping (Layer 1), may yield a more complete characterization of each patient’s epileptogenic network. Taken together, the results suggest that epileptogenic participation may be more appropriately conceptualized as sustained energy burden that can be triggered or activated by changes within the network, as has been demonstrated in neural fragility theory, rather than solely as relative deviation from baseline activity.

### 3.2 Normalization as a Structural Limitation, Not a Flaw

Baseline normalization allows ictal activity to be distinguished from the heterogeneous resting-state amplitudes that characterize clinical intracranial EEG, and methods built on this principle have been transformative for epilepsy surgery (Bartolomei et al., 2008; Bernabei et al., 2023). However, the same operation that enables robust ictal detection also attenuates a specific physiological signal: the contribution of tissue whose baseline is chronically elevated. The Hilbert decomposition results are consistent with a straightforward mechanism. Because the z-score denominator grows with baseline amplitude, a region that amplifies substantially during a seizure produces only a modest z-score when its resting baseline is high. The same logic applies to any framework that references ictal activity to a resting baseline (Epileptogenicity Index), to a resting network configuration (neural fragility), or to a discrete background rate (high-frequency oscillations). The dual-metric approach does not reject normalization; it supplies an independent absolute reference against which normalization can be cross-checked.

### 3.3 Relation to Existing Quantitative Methods

The dual-metric approach is complementary to established frameworks for characterizing epileptogenic networks (Bartolomei et al., 2017). The Epileptogenicity Index captures *relative activation*, how much a region departs from its own baseline spectrum (Bartolomei et al., 2008). Neural fragility captures *network instability*, how a region destabilizes a linear model of the intracranial network (Li et al., 2021). The Virtual Epileptic Patient framework simulates patient-specific seizure spread on a structural connectome to produce predictive models of seizure dynamics (Jirsa et al., 2017). High-frequency oscillation rates capture *transient events*, discrete fast-ripple phenomena counted against a background rate (Frauscher et al., 2017; Lisgaras et al., 2026). Dual-metric TKEO proposes an additional axis, *sustained absolute energy burden*, that may partially overlap with existing measures, particularly the Epileptogenicity Index which also derives from energy computation, but differs in its retention of absolute scale. The most informative characterization of an individual patient’s epileptogenic network is likely to require consideration of multiple complementary approaches. In clinical practice, EDNRs would be interpreted relative to the seizure onset zone and its tractography-derived propagation network, providing the surgeon with a second layer of network information that may identify targets not apparent from onset timing or structural connectivity alone. Notably, the present analysis uses the same ds003029 cohort on which neural fragility was originally validated (Li et al., 2021), providing a demonstration that a second, construct-independent analytical axis may surface network information not fully captured by the fragility framework on the same underlying recordings. A recent directional-connectivity analysis identified non-resected cortical regions assuming dominant directional roles in late ictal and postictal phases (Banappa et al., 2026), compatible with the energetic account that tissue energetically engaged throughout a seizure becomes progressively visible as the epileptogenic zone’s normalized dominance wanes. To check the rank-discordance operational definition against an independent benchmark, the Epileptogenicity Index was computed on the same cohort as a sanity check. EDNRs received high within-patient EI percentile (median 68.4) compared with non-involved tissue (median 44.4; two-tailed Mann-Whitney U, *p* = 2.5 × 10⁻⁶), confirming that EDNRs are not an artifact of the dual-metric definition: a methodologically independent quantitative framework also identifies them as pathologically involved. EI’s single scalar did not separate EDNR from PSOZ within the involved pool (AUC 0.45, 95% CI 0.38-0.51), as expected for a single-axis method asked to subdivide tissue along a dimension orthogonal to overall epileptogenicity. The cross-check is thus consistent with two complementary readings: the dual-metric framework and EI converge on the boundary between involved and non-involved tissue, while the dual-metric framework carries an additional axis that the present study characterizes.

### 3.4 Sampling Density as a First-Order Constraint on Network Characterization

The strong scaling of EDNR detection with electrode coverage (*r* = 0.899) and the absence of any plateau across the coverage range in these datasets may carry implications for how SEEG coverage is considered in network-level analyses. The dual-metric analysis cannot rescue information from electrodes that were never placed; no quantitative framework applied post hoc can compensate for insufficient sampling. Implantation strategies that prioritize minimal electrode coverage, whether for cost, time, or safety, may structurally under-sample the epileptogenic network independently of which analytical method is subsequently applied. This is not an argument for maximal implantation, which carries its own risks, but for recognizing that electrode strategy is upstream of every quantitative method and that adequate sampling density is a prerequisite for network-level inference from iEEG. The same observation constrains any cross-patient comparison on heterogeneous-coverage public data, including the descriptive clinical associations reported in this study.

### 3.5 Interpretation of the Descriptive Clinical Associations

The associations observed — higher EDNR counts and more frequent bilateral EDNRs in patients labeled not seizure-free — are compatible either with EDNRs marking tissue whose persistence in the post-operative brain contributes to seizure recurrence, or with EDNR burden marking more diffuse epileptogenic networks less amenable to focal intervention. The present data cannot distinguish these possibilities. The outcome column in both public datasets is a single-timepoint annotation without longitudinal surveillance, chart review, or knowledge of resection volume; for these reasons, the clinical associations are offered as descriptive observations only and no predictive claim is made. A study that could support predictive claims would require prospective standardized coverage, prospectively adjudicated resection boundaries, and longitudinal follow-up of sufficient duration, none of which are available in retrospective public data of this kind.

### 3.6 Biophysical Interpretation

The Hilbert decomposition results support a mechanistic account in which elevated baseline amplitude is the central feature of EDNRs and is sufficient to explain their attenuation under normalized metrics. This baseline elevation is consistent with chronic interictal hyperexcitability of the kind associated with the Rosenow-Lüders irritative zone (Rosenow and Luders, 2001) and with the long-recognized capacity of repeated focal activation to produce sustained local hyperexcitability through kindling-like mechanisms (Goddard et al., 1969). During seizures, EDNRs participate in seizure-frequency dynamics comparably to PSOZs and amplify substantially relative to non-involved tissue, indicating active ictal involvement rather than passive volume conduction. Whether the elevated baseline reflects chronic local hyperexcitability or dense local network connectivity cannot be fully disentangled in this dataset, but both mechanisms produce the same downstream effect: a large baseline scales the z-score denominator and attenuates the normalized expression of substantial absolute ictal energy. The same scaling logic applies to network-stability metrics that reference a resting system configuration and to event-based markers that count discrete transients against a baseline rate.

### 3.7 Limitations

The principal limitation of this study is the use of retrospective public iEEG data. The cohort was assembled from two OpenNeuro datasets with heterogeneous electrode implantation strategies, heterogeneous recording durations, and, critically, heterogeneous and single-timepoint outcome annotations. The outcome fields cannot support longitudinal inference, resection-volume analysis, or harmonized clinical endpoint scoring, and are used in this paper only for descriptive observations. Age and sex were unavailable for 4 of 56 patients in the analysis cohort, modestly limiting subgroup analysis. Sex- and gender-stratified analyses were not performed because the public datasets did not provide consistent stratification metadata across centers and the cohort size precluded reliable subgroup analysis; prospective cohorts should integrate sex- and gender-based analyses by design. The onset detection z-score threshold was adjusted per recording based on maximum z-score amplitude, which introduces a degree of parameter dependence in the rank computation; the amplitude-frequency decomposition results, which are the primary mechanistic findings, are independent of this threshold.

A second limitation concerns SEEG itself. Electrode contacts sample discrete points with effective sensing radii of approximately 5-10 mm (Buzsaki et al., 2012; Dubey and Ray, 2016), so regions of substantial seizure involvement may appear quiescent simply because no contact sits close enough to the generator. This raises an alternative source of rank discordance: a distant seizure generator could produce a signal whose distance-dependent attenuation preferentially reduces high-frequency content (Dubey and Ray, 2016) while preserving lower-frequency amplitude. The amplitude-frequency decomposition favors the hyperexcitability interpretation in most cases, because EDNRs show significantly elevated baseline amplitude that a pure volume-conduction account does not predict, but both mechanisms may coexist. Finally, the public datasets used here do not include diffusion tensor imaging or, for ds003029, SEEG electrode coordinates; the tractography component of Layer 1 therefore cannot be performed without patient-specific diffusion imaging.

A third limitation concerns the cohort-construction criterion itself. Six HUP recordings accessed through the iEEG.org portal were excluded from all analyses because they lacked entries in the ds004100 demographic table and therefore had no authoritative implant-modality field. The participants-table-based exclusion rule was retained to keep the cohort definition tied to the source-dataset metadata standard rather than to a downstream naming-convention heuristic.

### 3.8 Future Directions

The present findings motivate three distinct lines of prospective investigation. The first is direct: a prospective SEEG cohort with standardized implantation protocols, prospectively adjudicated resection volumes, and longitudinal surveillance of seizure outcome would directly test whether EDNR burden at specific anatomical sites, and in particular unresected EDNRs, carries clinical significance. Such studies are feasible at institutions running active SEEG programs and represent the natural next step beyond the descriptive analysis reported here. The second is integrative: multiple complementary axes for characterizing the epileptogenic network now exist, including baseline-normalized energy, network fragility, transient event markers, absolute energy burden, and directional connectivity dynamics (Banappa et al., 2026; Bartolomei et al., 2008; Frauscher et al., 2017; Li et al., 2021). Because these axes are largely orthogonal in construction, a joint framework that integrates several of them on paired datasets may outperform any single axis and clarify which combination of features is most informative in individual patients. The most complete characterization of a given patient’s seizure network is likely to require a tailored combination of these approaches rather than reliance on any one alone. The third is the completion of the two-layer framework proposed here: integration with patient-specific tractography to map the propagation network from identified seizure onset zones (Layer 1), enabling direct assessment of how EDNRs (Layer 2) relate spatially and structurally to the propagation network and whether that relationship modifies their clinical relevance. A fourth line is cross-modality extension: whether residual rank discordance between absolute and normalized scalp energy carries useful information beyond established source-localization (Plummer et al., 2019) and bipolar-montage approaches is an open empirical question, best addressed with simultaneous scalp-and-intracranial recordings in which intracranial EDNRs serve as ground truth.

## Conclusion

Dual-metric TKEO analysis characterizes energy-dominant network regions, a population whose contribution is attenuated under baseline-normalized metrics, and identifies elevated baseline amplitude as the distinguishing biophysical feature. These findings establish the second layer of a two-layer approach in which seizure onset detection and tractography-based propagation mapping constitute the first layer. Whether EDNRs correspond to regions along or beyond the propagation network, and whether unresected EDNRs carry significance for surgical outcome, define an agenda for prospective investigation in cohorts with patient-specific diffusion imaging and longitudinal follow-up.

## Ethical Statement

All intracranial EEG data analyzed in this study were obtained from publicly available, fully de-identified datasets distributed via OpenNeuro (ds004100 and ds003029) under their respective open data licenses. Because the data were public and de-identified, separate institutional review board approval was not required at the analyzing institution. The original data acquisition at the contributing centers was conducted under each center’s local institutional review board approval as documented in the source dataset publications.

## Data Availability

All intracranial EEG data used in this study are publicly available through OpenNeuro (datasets ds004100 and ds003029; https://openneuro.org). Single-timepoint outcome annotations are included in the dataset metadata (participants.tsv). The dual-metric TKEO method is described in sufficient detail in the Method section to enable independent implementation. Analysis code is not publicly released because it forms the basis of a pending US provisional patent application; requests for academic collaboration should be directed to the corresponding author.

## Acknowledgements

The author thanks the contributors of the publicly available datasets on OpenNeuro (ds004100 and ds003029) that made this research possible.

## Declaration of generative AI and AI-assisted technologies in the manuscript preparation process

During the preparation of this work the author used Claude (Anthropic) in order to assist with manuscript formatting, reference verification, and editorial revision of prose. After using this tool, the author reviewed and edited the content as needed and takes full responsibility for the content of the published article.

---

**Supplementary Figure S1.**
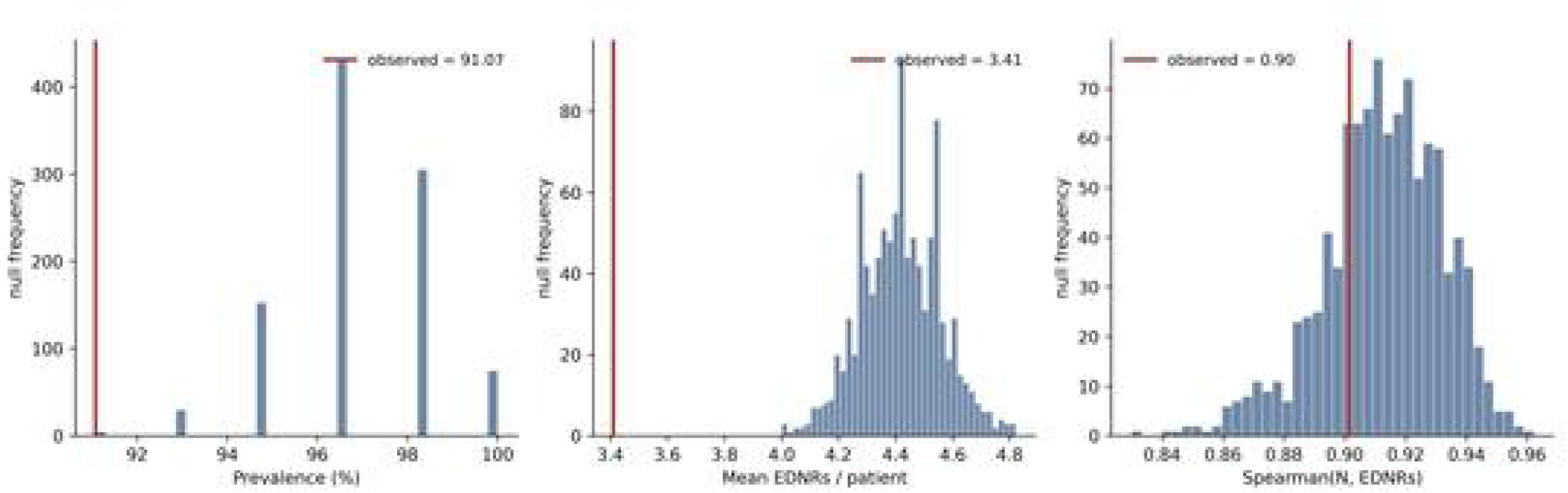

**Supplementary Figure S2.**
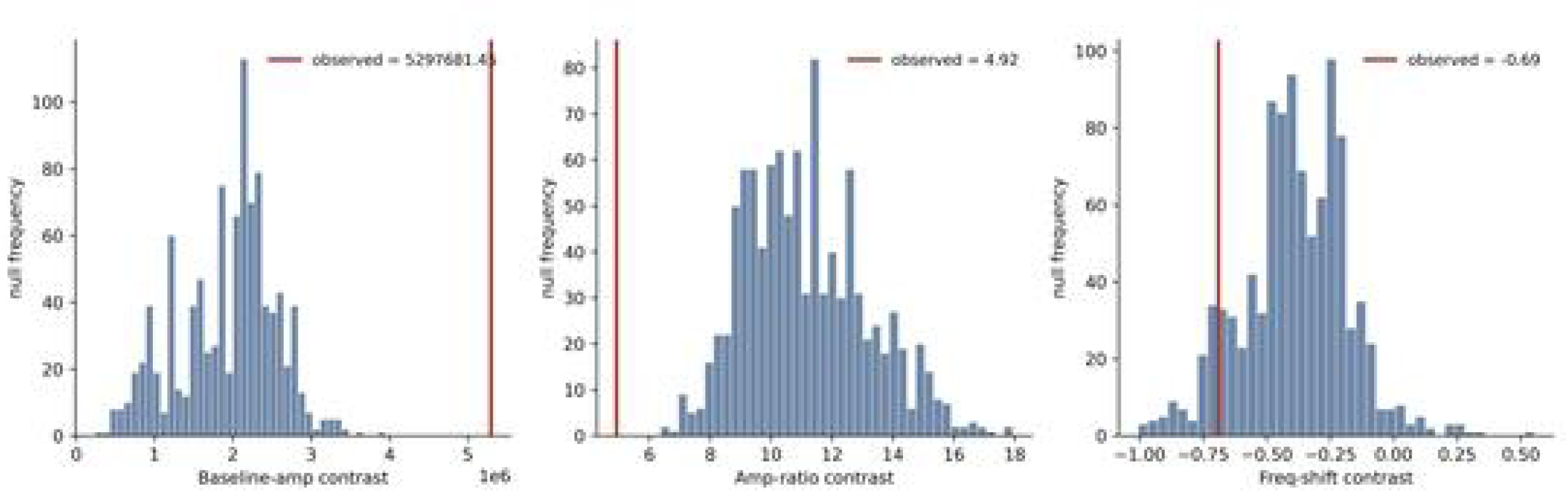

